# Digital Therapeutics Care Utilizing Genetic and Gut Microbiome Signals for the Management of Functional Gastrointestinal Disorders: Results from a Preliminary Retrospective Study

**DOI:** 10.1101/2021.10.01.21264214

**Authors:** Shreyas V Kumbhare, Patricia A Francis-Lyon, Dashyanng Kachru, Tejaswini Uday, Carmel Irudayanathan, Karthik M Muthukumar, Roshni R Ricchetti, Simitha Singh-Rambiritch, Juan A Ugalde, Parambir S Dulai, Daniel E Almonacid, Ranjan Sinha

**Affiliations:** Digbi Health, Mountain View, CA, USA; Health Informatics, University of San Francisco, San Francisco, CA, USA; C+, Research Center in Technologies for Society, School of Engineering, Universidad del Desarrollo, Santiago, Chile; Millennium Initiative for Collaborative Research on Bacterial Resistance, Santiago, Chile; Division of Gastroenterology, University of California San Diego, La Jolla, CA, USA

**Keywords:** multi-omic models, functional gastrointestinal disorders, IBS, diarrhea, constipation, digital therapeutics, non-pharmacological treatment

## Abstract

**Background:** Diet and lifestyle-related illnesses like obesity and functional gastrointestinal disorders (FGIDs) are rapidly emerging health issues worldwide. Research has focused on addressing FGIDs via in-person cognitive-behavioral therapies and lifestyle modifications focusing on diet modulation and pharmaceutical intervention. However, there is a paucity of research reporting on the effectiveness of digital care based on genome SNP and gut microbiome markers to guide lifestyle and dietary modulations on FGID associated symptoms and on modeling diseased groups or outcomes based on a combination of these markers.

**Objective:** This study sought to model subjects with FGID symptoms vs. those that do not present them, using demographic, genetic, and baseline microbiome data. Additionally, we aimed at modeling changes in FGID symptom severity of subjects at the time of achieving 5% or more of body weight loss in a digital therapeutics care program compared to baseline symptom severity.

**Methods:** A group of 177 adults with 5% or more weight loss on the Digbi Health personalized digital care program was retrospectively surveyed about changes in the symptomatology of their FGIDs and other comorbidities. The FGID subgroup rated their symptom severity on a scale of 1 to 5 at the beginning of the program and after successfully achieving >5% body weight decrease. During the intervention, personalized coaching for lifestyle changes, including diet and exercise, was delivered by both human and digital coaching. The demographic, genomic, and baseline microbiome data of the subgroup of participants (n=104) who self-reported any of six FGIDs (IBS, diarrhea, constipation, bloating, gassiness, and cramping) were compared with those who did not report FGIDs (n=73) and used as variables for a logistic model. The sum of reductions in symptom severity and IBS, diarrhea, and constipation symptom severity reduction were analyzed using the same variables in linear regression models.

**Results:** Gut microbiome taxa and demographics were the strongest predictors of FGID status. The digital therapeutics program implemented effectively reduced the summative severity of symptoms for 89.92% of users who reported FGIDs, with a highly significant reduction in severity (Wilcoxon signed-rank test, p=4.89e-17*). A mixture of genomic and microbiome predictors modeled the best reduction in summative FGID symptom severity and IBS symptom severity, whereas reduction in diarrhea symptom severity and constipation symptom severity were best modeled by microbiome predictors only.

**Conclusion:** A digital therapeutics program, informed by genomic SNPs and baseline gut microbiome and their interaction with participant diet and lifestyle, can effectively reduce functional bowel disorder symptomatology. While further research is needed for validation, demographics, microbiome taxa, and genetic markers can effectively inform models aiming at classifying subjects with FGIDs vs. those that do not have FGIDs and models assessing the reduction in symptom severity experienced by FGID sufferers. The methods and models presented here can readily be implemented to study other comorbidities where genetics and gut microbiome play a central role in disease etiology.

## Introduction

Diseases of the gastrointestinal tract affect 60-70 million people in the United States [1], and the total expenditure in 2015 for these illnesses was 135.9 billion dollars - greater than for other common diseases and likely to continue increasing. Abdominal pain alone accounted for $10.2 billion of the total costs [2]. GI afflictions and altered bowel habits affect the quality of life, social functioning and can result in considerable loss of productivity [3]. Early identification of appropriate markers for functional bowel disorders and a personalized therapeutic approach may help to reduce healthcare costs and improve quality of life.

Functional gastrointestinal disorders are conditions that present as normal upon examination of the GI system but still result in poor GI motility - primarily with symptoms in the middle or lower gastrointestinal tract [4]. These disorders include Irritable Bowel Syndrome (IBS), bloating, constipation, diarrhea, gas, and dyspepsia [4,5], among others. Although the pathophysiology of FGIDs is often complex due to their multifactorial nature, they are frequently encountered in both primary care and gastroenterology settings. FGIDs are thought to encompass bidirectional dysregulation of the gut-brain interaction via the gut-brain axis, gut microbial dysbiosis, visceral hypersensitivity, altered mucosal immune function, and abnormal gastrointestinal tract motility [6,7], and women appear to be afflicted at least twice as frequently as men [8].

Irritable Bowel Syndrome (IBS) is the most common functional bowel disorder and is prevalent in the range of 5–25% of the population and accounts for 36% of visits to a gastroenterologist. It is a gastrointestinal tract disease characterized by abdominal pain, altered bowel habits, and affected quality of life, but the absence of demonstrable organic disease is not considered life-threatening. IBS affects 10-15% of the US adult population [2,8]. Despite being a non-life-threatening disease, IBS can cause considerable pain and discomfort, and treatment often requires lifestyle changes and medication. The most common symptoms of IBS are chronic diarrhea - IBS-D (⅓ of IBS patients), chronic constipation - IBS-C, or both - IBS-M. Patients with IBS have a lower reported quality of life than sufferers of GERD and asthma [8].

Frequent and recurrent pain in the abdominal region not necessarily attributable to gut function is often referred to as Functional Abdominal Pain [14]. Children with functional abdominal pain were found to be significantly more likely to be obese [15]. FAP symptoms are not as common as other FGIDs and are not necessarily associated with food intake or passage but rather with psychiatric disorders. Management can involve psychotherapy and pharmaceutical interventions encompassing psychiatric regimens like anti-depressants, anticonvulsants, and treatments for other psychiatric disorders [16]. Functional bloating, on the other hand, is typically unlinked to psychiatric or organic causes [9]. Instead, it is a recurrent sensation of abdominal distention that may or may not be associated with measurable distention. Most people who report bloating have functional gastrointestinal disorders, and up to 96% of IBS patients report bloating. Bloating is 2x more commonly reported in women than men and typically worsens after meals [10,11]. Malabsorption of simple and complex carbohydrates and dietary fiber are commonly associated with both gas and bloating [6]. Gas is the byproduct of the bacterial fermentation of carbohydrates and protein in the large intestine, resulting in changes to the gut microbiome, increased short-chain fatty acids, and increased gas, diarrhea, abdominal pain, and bloating [9]. The exact connection between gas and bloating is not fully understood, but although intestinal gas may contribute to bloating, bloating does not necessarily result from more gas. Management often involves dietary modification, gut microbiome modulation, and lifestyle alteration [9].

Functional constipation is a functional bowel disorder that does not meet IBS criteria but presents as persistently difficult, infrequent, or seemingly incomplete defecation [10]. Chronic constipation occurs in up to 27% of people and affects all ages [10]. In women, constipation and associated straining to evacuate causes vaginal prolapse. Women undergoing prolapse surgery should understand the importance of correcting and managing constipation to decrease the risk of recurrence of the vaginal prolapse [12]. Depending on specific tests, constipation may be defined as (1) straining, hard stools, unproductive movements, infrequent stools, or incomplete evacuation; (2) less than three bowel movements per week OR daily stool weight less than 35 g/day OR straining for more than 25% of the period; (3) lengthy whole-gut or colonic transit. Surprisingly, stool frequency appears to have just a slender relationship with colonic transit and there are usually no demonstrable physiological abnormalities [10].

Functional diarrhea (FD) is a functional gastrointestinal disorder characterized by chronic or recurrent diarrhea not explained by structural or biochemical abnormalities of the gut. Functional diarrhea is characterized as the passage of loose or watery stools without abdominal pain or discomfort [10]. Unspecified diarrhea was reported by 4.8% of people throughout the US, but its duration and frequency are uncertain. Diarrhea is one of the most commonly reported symptoms for consulting a gastroenterologist and is also a common presenting issue among many patients in general practice [10]. Treatment of FD depends on establishing a correct diagnosis and needs to be distinguished from diarrhea-predominant irritable bowel syndrome (IBS-D) and other organic causes of chronic diarrhea. Once a physician has established the diagnosis, aggravating factors will need to be identified and eliminated, possibly including physiologic factors (e.g., small bowel bacterial overgrowth), psychological factors (e.g., stress and anxiety), and dietary factors (e.g., carbohydrate malabsorption) [13].

Diet is also considered an important trigger of gut-related symptoms. Poor nutrition, for example, consumption of highly processed and “fast” foods, has been implicated in FGID etiology [17,18]. Conversely, a Mediterranean diet has been associated with a lower prevalence of FGID [19]. A dietary therapeutics approach, for example a low-FODMAP diet in which rapidly fermentable carbohydrates that are poorly absorbed by the gut are eliminated or avoided as much as possible, is a typical dietary protocol for IBS patients [20]. Low-FODMAP diets have been associated with relieving other FGID symptoms, although functional dyspepsia seems least responsive to such a regimen [21,22].

Dietary fiber, which increases mucosal protein production, is widely used to treat chronic constipation [23]. This fiber is then digested in the colon, providing a substrate for microbial fermentation and resulting in byproducts such SCFAs, which have pro-motility effects and help with stool bulk and gas transit in the colon [24]. Also related to diet and FGID is the concept of non-celiac gluten sensitivity. Despite lacking serological and histological markers for celiac disease, a subset of FGID patients report significant alleviation of symptomatology upon elimination of dietary gluten and re-experience these symptoms upon reintroducing gluten [26].

Hormonal events, and genetic, environmental, and psychosocial factors have all been implicated to varying degrees in FGID etiology [10,20]. Sleep disturbance, dysfunctional coping, and psychological disturbance can correlate with symptom exacerbation. However, the nature of the link between psychosocial factors and FGIDs is poorly understood [10].

Both gut biome and genetics are likely contributors to FGID etiology. Evidence indicates a vital genetic component to FGIDs as demonstrated by prevalence within families and more substantial concordance between monozygotic versus dizygotic twins [27,28]. Functional dyspepsia has been associated with specific genotypes, including those associated with the ‘G-protein beta 3 subunit 825 CC genotype’, those associated with smooth muscles of the GI tract, and others associated with abdominal pain [29,30]. One evidence of a mutation linked to IBS was replicated across two studies, and although the mutation is found in only approximately 2% of IBS patients, this finding indicates the influence genetics may have on IBS symptoms [31,32]. Functional constipation has also been associated with specific genes [30]. Much of the research around genome-wide association studies and FGIDs is hampered by small sample sizes; however, the study of the genetics of FGID is rapidly evolving. The gut biome is also extensively implicated in FGID and particularly IBS pathogenesis [34,35]. Both IBS and functional dyspepsia have been shown to arise in susceptible individuals following a course of acute onset gastroenteritis [36]. Recent studies have revealed dysbiosis of the gut microbiota in constipated patients compared with healthy controls, associated with suppressed intestinal motility by metabolites produced by intestinal bacteria [24,37]. Other research has elucidated several mechanisms playing important roles in IBS. A dysregulated gut-brain axis has been adopted as a suitable model for IBS, and poor gut microbiome diversity may contribute to the onset and exacerbation of IBS symptoms. Dietary fibre appears to influence the gut microbiota, encouraging the growth of beneficial probiotics while preventing pathogenic and obesogenic bacteria from overgrowing [24,25]. Although clinical trials, which have attempted to characterise the gut microbiota in IBS, do not yet allow for a causal role to be inferred, they do confirm alterations in both community stability and diversity [38]. Although large-scale genomic analyses of IBS patients are lacking [20], numerous candidate genes have been potentially implicated in IBS. However, most studies involve small sample sets, have not been independently replicated, or are otherwise not robust. What seems clear so far is that most IBS sufferers either share several common gene variants that each nominally contribute toward the overall risk of the disease, or, for a subset of sufferers, a few highly penetrant alleles are likely the significant risk factors. Given that IBS spans both complex polygenic conditions and rare single-gene forms, it evidences the need for different strategies to identify these genetic factors.

Our aim for this study was to assess how effective a digital therapeutics intervention personalized on genomic SNPs and gut microbiome signals were at reducing symptomatology of FGIDs. Additionally, we aimed at building statistical models to describe the impact of each predictor (demographics, genomic SNPs, and gut microbiome) on the likelihood of a subject presenting or not FGIDs, and then model reduction of summative symptom severity (IBS, diarrhea, constipation, bloating, gas, and abdominal pain) as well as reduction of symptom severity in IBS, diarrhea, and constipation.

## Material and Methods

### Subject enrollment

Subjects were recruited from those who achieved 5% or more body weight loss when enrolled in the Digbi Health personalized digital care program, which delivers both human and digital personalized coaching for lifestyle changes, including diet and exercise, informed by demographics, genome SNP, and gut microbiome data (See supplementary methods for details). Only those subjects who had retrospectively responded to questions about symptomatology of their FGIDs and other comorbidities at the start of the program and after successful weight loss were included in the study (Figure 1). The average number of days for participants in the program at the time of the survey was 84.26 days. The final cohort studied in this manuscript included 177 subjects with either genome SNP (n=169) or baseline gut biome (n=168) or both analyses performed. Subjects were divided into two groups for comparisons and models: those who reported any of 6 FGIDs (IBS, diarrhea, constipation, bloating, gassiness, and cramping) at baseline or at the time of survey (n = 104), and those who reported no FGIDs (n=73).

**Figure 1.**
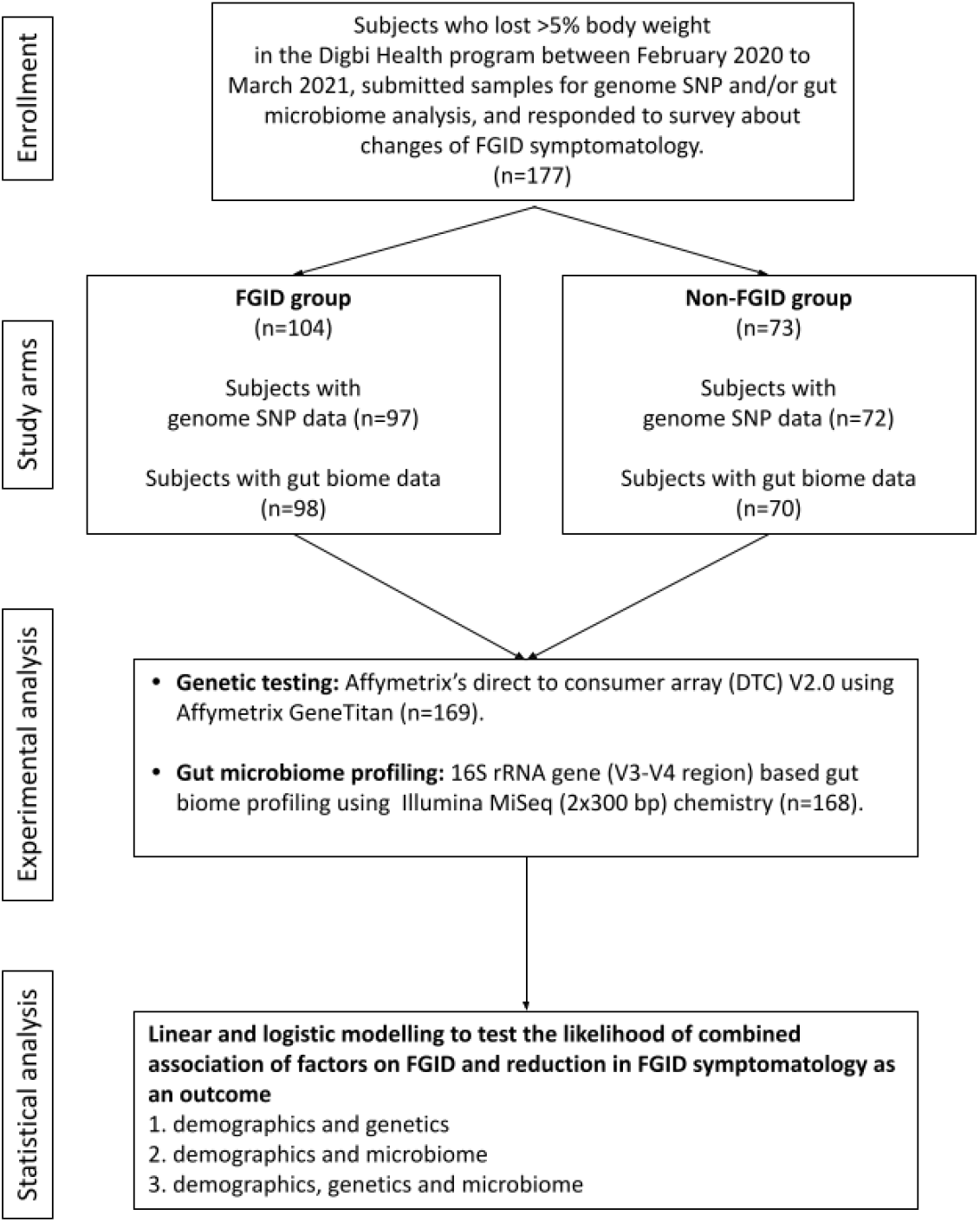
Study design flowchart.

### Sample collection and processing: Genome SNP array and gut microbiome profiling

Subjects self-collected saliva samples using buccal swabs (Mawi Technologies iSwab DNA collection kit, Model no. ISWAB-DNA-1200) and fecal samples using fecal swabs (Mawi Technologies iSWAB Microbiome collection kit, Model no. ISWAB-MBF-1200). Sample collection was completed by following standardized directions provided to all subjects in an instruction manual. Saliva DNA extraction, purification, and genotyping using Affymetrix’s Direct to Consumer Array version 2.0 (“DTC”) on the Affymetrix GeneTitan platform was performed by Akesogen Laboratories in Atlanta, GA. Sample processing of baseline (pre-intervention) fecal samples was followed by the 16S rRNA gene amplicon sequencing also performed at Akesogen Laboratories in Atlanta, GA. DNA extraction was performed using Qiagen MagAttract Power Microbiome DNA Kit on an automated liquid handling DNA extraction instrument. The V3-V4 region of the 16S rRNA gene was amplified and sequenced on the Illumina MiSeq platform using 2 × 300 bp paired-end sequencing [39]. Sequence reads were demultiplexed, and ASVs generated using DADA2 in QIIME2 (version 2020.8) [40]. We trim primers off the reads and low-quality bases (Q <30). Taxonomic annotation was performed using the Naive Bayes classifier against the 99% non-redundant Silva 138 reference database [41]. We excluded hits to Mitochondria, Chloroplast, Eukaryota, and unassigned taxa at the phylum level.

### Statistical analysis

Survey data from 177 respondents (with either genome SNP or gut biome data) over the course of their successful weight loss journey in the Digbi Health personalized digital care program were analyzed retrospectively. Variables included demographic (gender, age, weight, and BMI during gut microbiome sample collection and weight loss achieved during the program), genomic SNPs, and baseline gut microbiome data. We used the Wilcoxon sum rank/signed-rank test or Kendall correlation test, as appropriate, to assess: (a) the change in the severity of either of the FGID symptoms and a summative severity change and (b) the effect of variables on the change in severity. Significance results were adjusted for multiple comparisons using the false discovery rate (FDR) correction method for the microbiome data.

### Genome SNP related statistics

The 16 SNP predictors, associated with lactose intolerance, gluten sensitivity, milk and peanut allergies, caffeine metabolism, and inflammatory markers (TNF & IL10), were from Digbi Health curated panels used for personalized interventions for subjects (Table S1). Each SNP value was encoded as the number of risk alleles (0, 1, or 2) for each subject.

### Microbiome related statistics

Bacterial genera abundances were analysed for 168 survey respondents with baseline gut microbiome data available, using Qiime276, qiime2R [42], and phyloseq [43]. The following microbial features were filtered out from downstream analysis: (a) ASVs not classified at the phylum level, (b) phyla that had <25 ASVs (*Elusimicrobiota, Nanoarchaeota, Bdellovibrionota, WPS-2*), (c) uncultured and Incertae Sedis taxa, (d) genera that had < 30 reads in at least 15% of samples, and (e) genera for which > 25% of samples had zero read count. In total, 105 genera were kept in downstream analysis. The abundance of these bacterial genera was transformed to centered log-ratio (CLR) using the zCompositions package [44] after first replacing zeros with pseudo counts based on a Bayesian-multiplicative replacement from the zCompositions package [45]. Permutational multivariate analysis of variance (PERMANOVA) was performed on the gut microbiome Aitchison distance matrix, using gender, BMI (closest to date of gut biome sampling), and FGID status as variables using the CLR transformed abundances. Additionally, the read counts after zero replacement were transformed using additive log-ratio (ALR) to be utilized in downstream model statistics (see below) [46–48].

### Model statistics

Linear and logistic regression models were built and visualizations generated using the R stats [49], ggplot2 [50], pscl, car, pROC, Metrics, caret, glmnet, tidyverse, lubridate, imputeTS, and ggpubr packages. In order to utilize lasso regression for variable selection before fitting linear and logistic models, SNPs with >10% missing values were removed, then remaining missing SNPs were imputed to their most frequent value (mode). This resulted in the removal of rs4713586 gluten sensitivity SNP from both reductions in summative symptom severity and reduction in constipation symptom severity models. In order to avoid poor performance in regression models, variables with Pearson correlation to another variable of >=80% were removed. This excluded two SNPs from all regression models that incorporated DNA: rs182549 of the lactose persistence haplotype was removed while the more highly cited correlated (Pearson, 0.99) haplotype SNP, rs4988235, was retained. Additionally, IL10 SNP rs3024496 was removed as being highly correlated (Pearson, -0.95) with rs1800896, while another IL10 SNP was retained as having a higher risk in the population [51].

For regression models, we transformed bacterial abundance data using the additive log-ratio (ALR) [46, 48], which maintains subcompostional coherence, permitting genera to be removed in downstream analysis (for example, removing insignificant predictors from a regression model). For this, the easyCoda package [52] was employed to analyse all 105 microbial genera utilizing variances, variances explained, and Procrustes correlations to select candidate references for ALR according to methodology of Greenacre *et al* [53]. An additional criterion we employed was prevalence, as zero values are problematic with log-ratios. Based on these criteria, *Blautia* was the selected reference. In all cases of highly correlated microbe pairs, the unclassified microbe was removed, or if both were unclassified, then the microbe with the larger mean absolute correlation in that dataset was removed. This resulted in removing up to 5 microbes (of 105) from regression models incorporating microbial predictors. Gender and SNPs were scaled to a range between -1 and 1. Age and gender were the two demographic variables used in all models and were used with no transformation. All predictors were standardized.

After the above data preparation, lasso was employed, retaining predictors with non-zero coefficients with optimal lambda chosen by a 5-fold cross-validation grid search. The lambda resulting in the minimum mean cross-validated error was selected, or if this resulted in a paucity of predictors, then a plateau lambda in error vs. lambda plot having a sufficient number of predictors was selected. Mean squared error was employed in linear lasso regression, while the mean absolute error was used in logistic regression models. After lasso, the step function (stats package) was employed, using the Akaike information criterion (AIC) to obtain a high-quality fit. If any insignificant variables remained, these were removed from the least significant variable resulting in a final interpretable model for each investigation and set of predictors.

After pre-processing, modeling of FGID vs. non-FGID was conducted by fitting a logistic regression model to demographic and 14 SNPs genomic data (D+G model), producing coefficients to describe the impact of each predictor on FGID in this cohort. A second logistic model was fit with demographic plus 104 baseline gut microbiome genera remaining after pre-processing (D+M model)). A third logistic model similarly employed lasso for variable selection from demographic variables, 14 genomic SNPs, and 104 baseline genera (D+G+M model). As described above, the step function was employed for all three logistic models, followed by removing any remaining insignificant variables, resulting in the final interpretable models.

For those respondents who self-reported any of the 6 FGIDs, change in symptom severity was analysed with respect to their demographic, genome SNPs, and baseline gut microbiome data. 104 survey respondents rated the severity of their FGID symptoms on a scale of 1 to 5. A linear regression model was fit to describe the change in summative symptom severity as a function of demographic and genomic variables (D+G model). As above, this model was compared with two additional models: demographic plus microbial predictors (D+M model) and demographic, genomic, plus microbial predictors (D+G+M model). Lasso regression was employed as above for variable selection, followed by the use of the step function. Additionally, changes in IBS, diarrhea and constipation symptom severity were modeled using linear regression as above with D+G, D+M, and D+G+M predictors.

## Results

### 1. Subject characteristics

In total, 177 subjects who were successful at losing 5% or more body weight and had genetic and/or gut biome data while enrolled in the Digbi Health program were surveyed to assess any changes in the symptomatology of their FGIDs (Figure 1). We compared baseline characteristics of those who reported FGIDs vs. those who did not. The distributions of gender, age, and BMI are seen in Table 1. In this dataset, a significant difference was found in gender (χ^2^ test, p=0.004*) and initial BMI (Wilcoxon sum rank test, p=0.009*), between those who reported FGID and those who did not, but no significant difference was found in age (Wilcoxon sum rank test, p=0.60). Subsequently, we investigated the effect of gender, BMI, and FGID status on the beta diversity of the baseline gut microbiome of subjects. The PERMANOVA analysis on Table S2 shows that gender had a significant effect on the beta diversity (R^2^=0.012, p=0.030*), whereas BMI (R^2^=0.007, p=0.279) and FGID status (R^2^=0.005, p=0.532) did not.

**Table 1.** Distribution and demographics of FGID and Non-FGID groups in the study.

| Descriptor variable | FGID group<br>(n=104) | Non-FGID<br>group (n=73) | P-value |
| --- | --- | --- | --- |
| **Gender, n (%)** |  |  |  |
| **Male** | 12 (11.54%) | 22 (30.14%) | <0.01* |
| **Female** | 92 (88.46%) | 51 (69.86%) |  |
| **Age, in years**<br>**(Median ± SD)** | 49 ± 11.92 | 51 ± 12.25 | 0.60 |
| **BMI closest to gut**<br>**biome sampling**<br>**(Median ± SD)** | 31.17 ± 7.37 | 33.51 ± 7.50 | <0.01* |

### 2. Gut microbiome taxa are a stronger predictor of FGID than the genomic SNPs analyzed in this study

SNP values were not significantly different (Welch’s two-sample t-tests, results not shown) between respondents with FGID and those without. Logistic regression modeled the associations of demographic, genome SNP, and baseline gut microbiome variables with FGID status in this cohort (Tables 2, 3, and 4), fitting separate average effect size for each predictor while controlling for all other model variables. The D+G model described females in this cohort as 3.26 times more likely than males to suffer FGID while controlling for the genomic predictors in the model (Table 2), and each risk allele of the rs2187668 gene was associated with a 2.93 times greater likelihood of being an FGID sufferer. Similarly, risk alleles for rs2472297 and rs9275596 were associated with the lowered likelihood of FGID - conferring 0.45 and 0.56 times likelihood of being an IBD sufferer.

**Table 2.** FGID vs Non-FGID Logistic Model: Demographics + Genomics (D+G)

| Variable | OR | 2.5% C.I. | 97.5% C.I. |
| --- | --- | --- | --- |
| Gender | 3.255 | 1.421 | 7.856 |
| Caffeine Metabolism (rs2472297), Risk Allele C | 0.447 | 0.211 | 0.888 |
| Gluten Sensitivity (rs2187668), Risk Allele T | 2.926 | 1.140 | 8.269 |
| Peanut Allergy (rs9275596), Risk Allele C | 0.556 | 0.323 | 0.935 |

McFadden pseudo R<sup>2</sup>: 0.089

**Table 3.** FGID vs Non-FGID Logistic Model: Demographics + Microbiome (D+M)

| Variable | OR | 2.5 % C.I. | 97.5 % C.I. |
| --- | --- | --- | --- |
| Gender | 2.334 | 1.213 | 4.686 |
| *Ruminococcus torques* group | 1.193 | 1.039 | 1.391 |
| *Akkermansia* | 1.076 | 1.011 | 1.151 |
| *Holdemanella* | 0.932 | 0.878 | 0.986 |
| Unclassified genus CAG-56 of *Lachnospiraceae* family | 1.095 | 1.024 | 1.175 |
| Unclassified genus UCG-010 of *Oscillospirales* order | 0.904 | 0.839 | 0.970 |
| *Anaerostipes* | 0.806 | 0.680 | 0.938 |
| *Haemophilus* | 1.115 | 1.034 | 1.210 |
| *Fusicatenibacter* | 0.894 | 0.803 | 0.985 |
| *Terrisporobacter* | 1.076 | 1.008 | 1.153 |

McFadden pseudo R<sup>2</sup>: 0.220

**Table 4.**
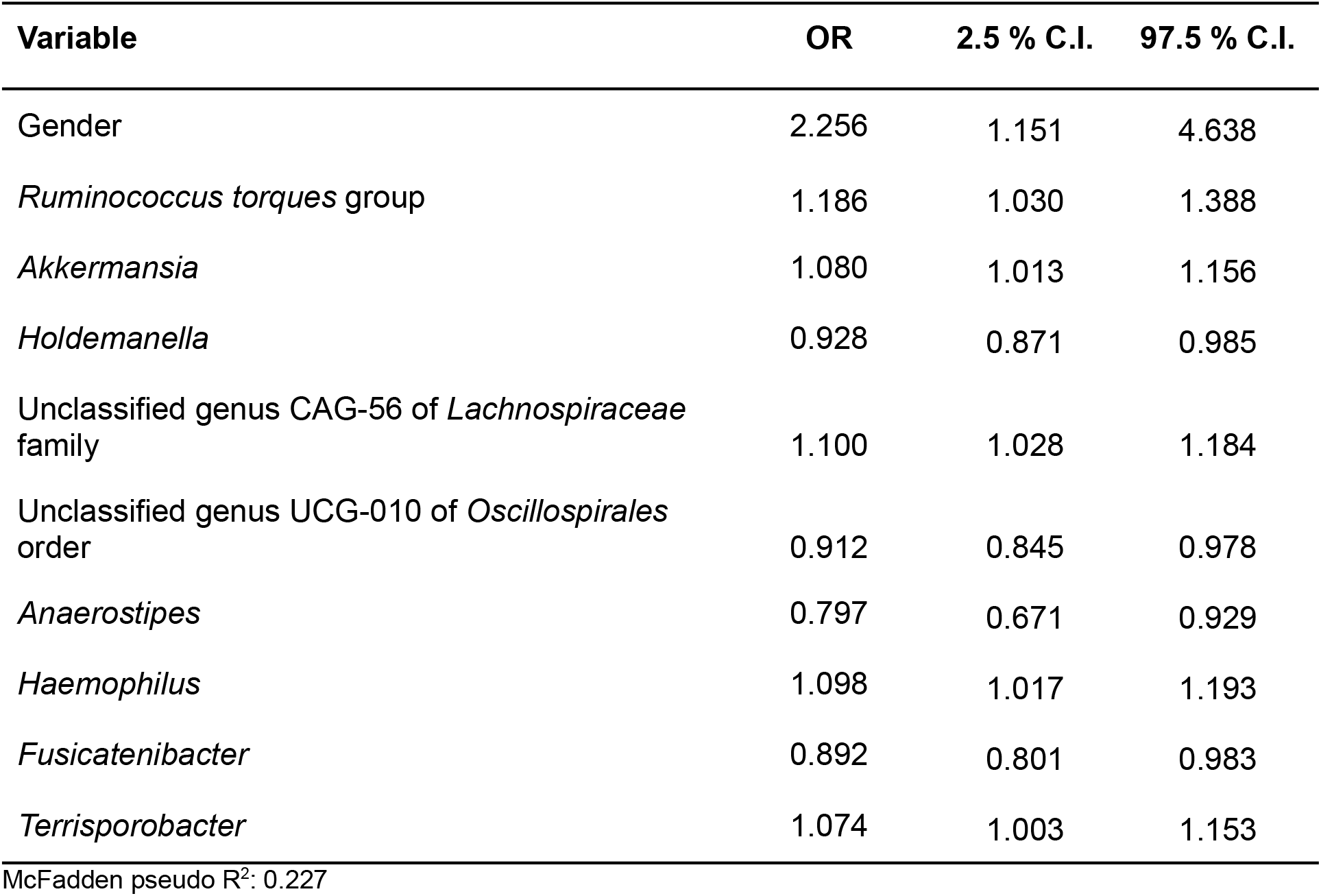
FGID vs Non-FGID Logistic Model: Demographics + Genomics + Microbiome (D+G+M)

| Variable | OR | 2.5 % C.I. | 97.5 % C.I. |
| --- | --- | --- | --- |
| Gender | 2.256 | 1.151 | 4.638 |
| *Ruminococcus torques* group | 1.186 | 1.030 | 1.388 |
| *Akkermansia* | 1.080 | 1.013 | 1.156 |
| *Holdemanella* | 0.928 | 0.871 | 0.985 |
| Unclassified genus CAG-56 of *Lachnospiraceae* family | 1.100 | 1.028 | 1.184 |
| Unclassified genus UCG-010 of *Oscillospirales* order | 0.912 | 0.845 | 0.978 |
| *Anaerostipes* | 0.797 | 0.671 | 0.929 |
| *Haemophilus* | 1.098 | 1.017 | 1.193 |
| *Fusicatenibacter* | 0.892 | 0.801 | 0.983 |
| *Terrisporobacter* | 1.074 | 1.003 | 1.153 |

McFadden pseudo R<sup>2</sup>: 0.227

In a second logistic model, we studied the associations of D+M together on the likelihood of a subject having an FGID (Table 3). The nine taxa identified by lasso regression were used as predictors for a logistic regression model to describe the classification of 168 subjects into their corresponding FGID status, using the genus *Blautia* as the reference denominator for alr. In this model, the effect of the female gender while controlling for microbial predictors was an average 2.33 times likelihood of FGID compared with the male gender. Genera *Ruminococcus* torques group, *Akkermansia*, unclassified genus CAG-56 of *Lachnospiraceae* family, *Haemophilus*, and *Terrisporobacter* were all associated with increased FGID. Whereas *Holdemanella*, unclassified genus UCG-010 of *Oscillospirales* order, *Anaerostipes*, and *Fusicatenibacter* were all associated with decreased (protective of) FGID status.

In a third logistic model studying the associations of D+G+M variables together on the likelihood of a subject having FGID status (Table 4), no SNPs had a significant association with FGID risk. Variables in Tables 3 and 4 are identical, with just slight variations in odd ratios. Not surprisingly, pseudo R2 values from the D+M model (0.220) and the D+G+M model (0.227) were very similar, but most importantly, improved from that of the D+G model (0.089).

### 3. Subjects reported a reduction in severity of FGID related symptoms over the course of treatment

The proportion of subjects who experienced at least one point improvement in symptom severity ranged from 75.32% for diarrhea to 90.63% for bloating (Figure 2). Improvement in summative severity across the 6 FGIDs was seen by 89.42% of respondents, with an average summative reduction of 51.17% (Wilcoxon signed-rank test, p=4.89e-17*). The improvement in FGID symptomatology over the course of digital therapeutics intervention (percent summative reduction) was not correlated with percent weight loss (Kendall = 0.12, p=0.91), age (Kendall = -0.66, p=0.51), or gender (Wilcoxon sum rank, p=0.809).

**Figure 2.**
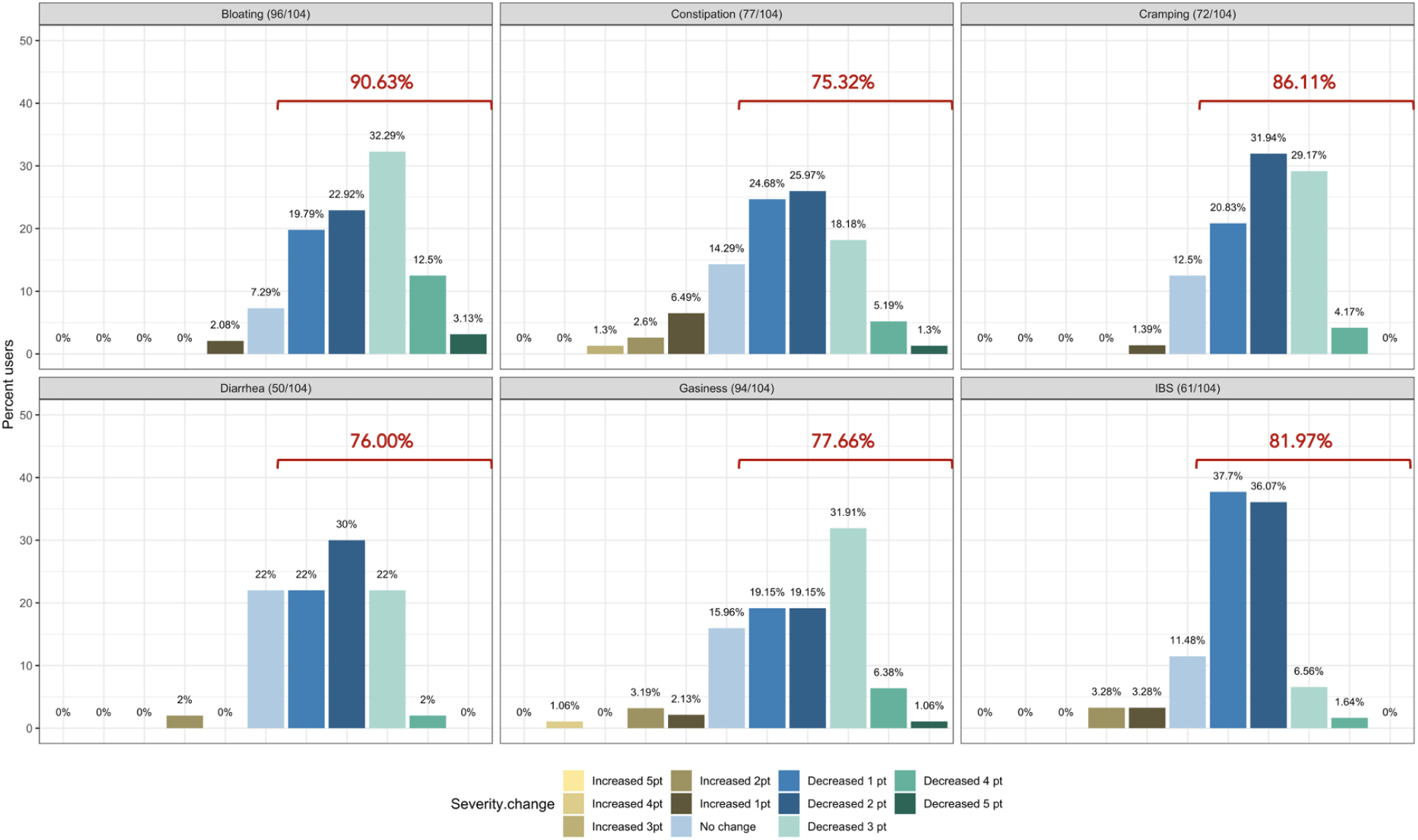
Self-reported symptom severity change (number of levels increased/decreased severity) in functional gastrointestinal disorders from baseline to time of survey.

Individually, we noted an average 45.93% reduction in the severity of IBS (Wilcoxon signed-rank, p=1.61e-08*), 61.01% average reduction in the severity of bloating (Wilcoxon signed-rank test, p=2.97e-16*), 38.55% average reduction in the severity of gassiness (Wilcoxon signed-rank, p=6.33e-12*), 61.69% average reduction in the severity of cramping/belly pain (Wilcoxon signed-rank, p=4.33e-12*), 37.54% average reduction in the severity of constipation (Wilcoxon signed-rank, p=3e-09*) and a 48.97% average reduction in the severity of diarrhea (Wilcoxon signed-rank, p=1.65e-07*).

### 4. Genomic and microbiome predictors can model reduction in summative FGID symptom severity as a result of the digital therapeutics intervention

In a linear D+G model of reduction in summative FGID symptom severity (Table S3), each additional risk allele for SNPs rs4639334 and rs7775228 was associated with an increase in self-reported summative FGID symptom severity.

In a linear D+M model of reduction in summative FGID symptom severity (Table S4), gut microbial genera Candidatus *Soleaferrea, Eubacterium hallii* group, *Alistipes*, and *Desulfovibrio* were all associated with an increase in self-reported summative FGID symptom severity. Microbial genera *Ruminococcus torques* group, *Intestinimonas*, unclassified genus GCA-900066575 of *Lachnospiraceae* family, and *Megasphaera* were all associated with a self-reported reduction in summative FBD symptom severity.

In a linear D+G+M model of reported FGID symptom severity (Table 5), risk alleles of the same two SNPs identified in Table S3 were again associated with an increase in self-reported summative FGID symptom severity. Similarly, gut microbial genera *Desulfovibrio*, Candidatus *Soleaferrea*, and *Eubacterium ventriosum* group Microbial genera *Megasphaera*, unclassified genus CAG-352 of *Rumniococcaceae* family, *Ruminococcus torques* group, *Streptococcus*, and *Intestinimonas* were all associated with a self-reported reduction in summative FGID symptom severity. Adjusted R^2^ values were 0.124 for the D+G model, 0.318 for the D+M model, and 0.442 for the D+G+M model. This indicates that the fit of the models improved when adding microbiome predictors, with the best fit model for a reduction in summative FGID symptom severity containing a mixture of genomic SNP and microbiome variables.

**Table 5.** Reduction in Summative FGID Symptom Severity Linear Model: Demographics + Genomics + Microbiome (D+G+M)

| Variable | Estimate | Std. Error | t value | Pr(>\|z\|) |
| --- | --- | --- | --- | --- |
| *Megasphaera* | 0.495 | 0.136 | 3.631 | <0.001* |
| *Desulfovibrio* | -0.517 | 0.105 | -4.910 | <0.001* |
| Unclassified genus CAG-352 of *Rumniococcaceae* family | 0.257 | 0.104 | 2.477 | 0.015* |
| Gluten Sensitivity (rs4639334), Risk Allele A | -2.013 | 0.792 | -2.541 | 0.013* |
| *Ruminococcus torques* group | 0.646 | 0.246 | 2.626 | 0.010* |
| Gluten Sensitivity (rs7775228), Risk Allele C | -1.936 | 0.828 | -2.338 | 0.022* |
| *Streptococcus* | 0.505 | 0.218 | 2.322 | 0.023* |
| *Intestinimonas* | 0.319 | 0.116 | 2.756 | 0.007* |
| *Soleaferrea* | -0.461 | 0.150 | -3.076 | 0.003* |
| *Eubacterium ventriosum* group | -0.290 | 0.134 | -2.157 | 0.034* |

Adjusted $R^2$ : 0.442

### 5. In the descriptive models, reduction in IBS symptom severity was better explained by a mixture of genomic and microbiome predictors, whereas reduction in diarrhea and constipation symptom severity was better explained by microbiome predictors only

Tables 6, 7, and 8 present linear D+G+M models for reduction of symptom severity for IBS, constipation, and diarrhea, respectively. Tables S5, S7, and S9 present linear D+G models, and Tables S6, S8, and S10 present linear D+M models for reduction of symptom severity for IBS, constipation, and diarrhea, respectively. Adjusted R^2^ values for the reduction in IBS symptom severity models were 0.130 for the D+G model, 0.432 for the D+M model, and 0.487 for the D+G+M model. Adjusted R^2^ values for the reduction in constipation symptom severity models were 0.038 for the D+G model, 0.413 for the D+M model, and 0.389 for the D+G+M model. Adjusted R^2^ values for the reduction in diarrhea symptom severity models were 0.090 for the D+G model, 0.610 for the D+M model and 0.528 for the D+G+M model. This shows that genomic SNP models performed relatively poorly and that the inclusion of microbiome variables constantly improved the fit of the models. Interestingly, the D+G+M was the best fit model for IBS symptom reduction, whereas, for constipation and diarrhea symptom reduction, the D+M models were the best fit.

**Table 6.** Reduction in IBS Symptom Severity Linear Model: Demographics + Genomics + Microbiome (D+G+M)

| Variable | Estimate | Std. Error | t value | Pr(>\|z\|) |
| --- | --- | --- | --- | --- |
| Gluten Sensitivity (rs7775228), Risk Allele C | -0.566 | 0.225 | -2.515 | 0.016* |
| *Unclassified genus Clostridia UCG-014* | 0.092 | 0.024 | 3.844 | <0.001* |
| *Escherichia-Shigella* | 0.087 | 0.028 | 3.122 | 0.003* |
| *Fusicatenibacter* | -0.096 | 0.038 | -2.501 | 0.016* |
| *Megasphaera* | 0.101 | 0.034 | 3.007 | 0.004* |
| *Moryella* | -0.079 | 0.028 | -2.775 | 0.008* |

Adjusted R<sup>2</sup>: 0.487

**Table 7.** Reduction in Constipation Symptom Severity Linear Model: Demographics + Genomics + Microbiome (D+G+M)

| Variable | Estimate | Std. Error | t value | Pr(>\|z\|) |
| --- | --- | --- | --- | --- |
| *Parabacteroides* | 0.231 | 0.073 | 3.174 | 0.003* |
| *Eubacterium coprostanoligenes* group | -0.161 | 0.051 | -3.125 | 0.003* |
| *Lachnospira* | 0.123 | 0.047 | 2.606 | 0.013* |
| Gluten Sensitivity (rs7775228), Risk Allele C | 0.757 | 0.327 | 2.311 | 0.026* |

Adjusted R<sup>2</sup>: 0.389

**Table 8.**
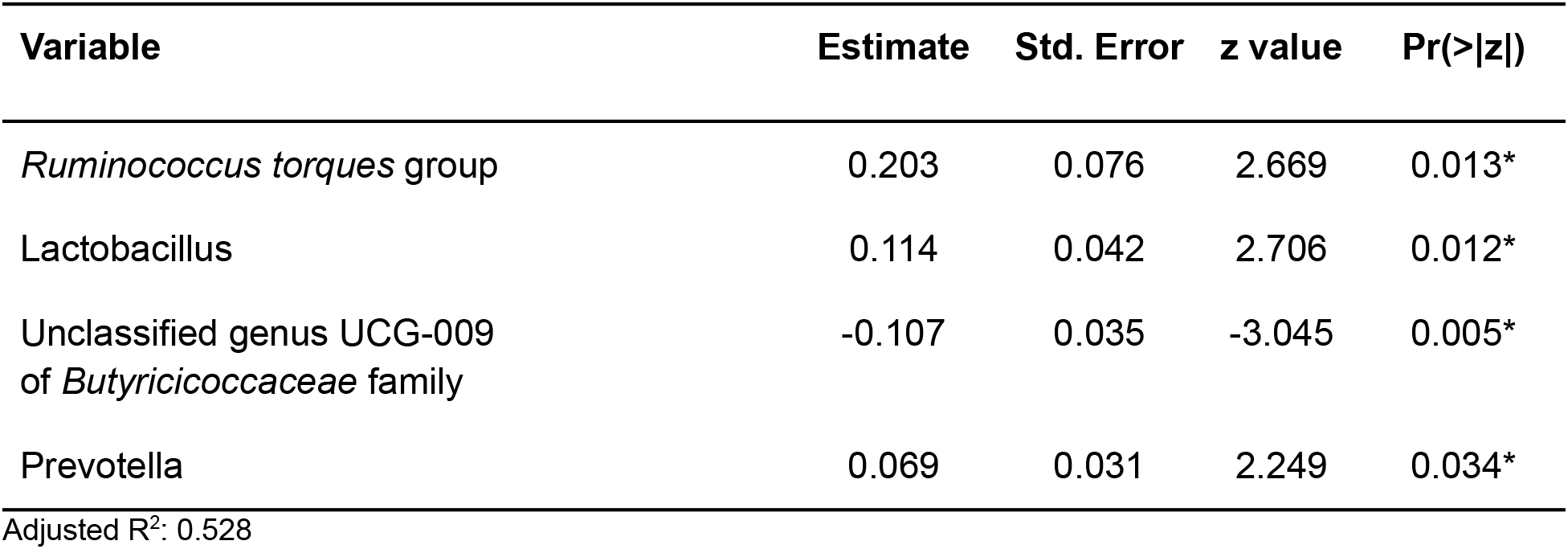
Reduction in Diarrhea Symptom Severity Linear Model: Demographics + Genomics + Microbiome (D+G+M)

Looking at the best fit models, *Unclassified genus Clostridia UCG-014, Escherichia-Shigella*, and *Megasphaera* were associated with reduction of IBS symptom severity, whereas Moryella, Fusicantenibacter, and risk alleles of rs7775228 (gluten sensitivity) were associated with an increase in For constipation, there were only microbial taxa in the D+M model: *Parabacteroides*, Unclassified genus of *Anaerovoracaceae* Family XIII AD3011 group, *Lachnospira and Terrisporobacter were* associated with a reduction in symptom severity, whereas *Eubacterium coprostanoligenes* group was associated with the increase. For diarrhea change in symptom severity and constipation, only microbial taxa were found significant in the D+M model: *Intestinimonas, Prevotella, Lactobacillus*, and *Phascolarctobacterium* were associated with a reduction in symptom severity, while Unclassified genus UCG-009 of the *Butyricicoccaceae* family was associated with the increase.

## Discussion

Out of 177 subjects enrolled in this study who successfully lost 5% or more body weight through a digital therapeutics program, 104 presented one or more FGIDs. These FGID sufferers were significantly different from the non-FGID group in terms of gender and BMI at the time of sampling (Table 1). Additionally, gender was significantly associated with the composition of baseline gut microbiome samples in these subjects (Table S2). Thus, using gender as a demographic variable, we trained logistic regression models to differentiate FGID status using D+G, D+M, and D+G+M variables. The models confirmed that the female gender was associated with a higher prevalence of FGID as seen in the logistic regression D+G model, where females were 3.26 times more likely to be FGID sufferers than males while holding genomic predictors constant (Table 2). This is in accordance with what has been reported elsewhere [54]. Additionally, when baseline microbiome was added to the model (D+G+M model), females were on average 2.26 times more likely to suffer from FGID than males while holding constant both genomic and microbial predictors (Table 4). Thus, some of the gender association in the D+G model is explained by microbiome variables in the D+G+M model, reinforcing the role of gender in shaping the baseline gut microbiome of subjects [55].

Of note, the SNP: gluten sensitivity (rs2187668, risk allele T) was seen to be strongly associated with FGID status in our cohort. This SNP variant is identified as HLA-DQ2.5 and has been reported as one of the most common HLA-DQ2 haplotypes associated with celiac disease [56]. Interestingly, the D+G+M model did not select any genomic variables and was identical to the D+M model (same variables but slightly different odd ratios), and not surprisingly, the pseudo R^2^ scores for these models are similar (0.227 for D+G+M vs. 0.220 for D+M). These models have pseudo R^2^ scores higher than for the D+G model (0.089), indicating that baseline microbiome better-classified participants having FGID than models based on genomic predictors and that the addition of SNPs did not improve classification of FGID by gender plus baseline microbiome. Many of the microbiome taxa variables identified in the models have already been reported in the literature associated with FGIDs. Previous studies show a strong association of the *Ruminococcus torques* group with FGIDs [57].

We then investigated the change in reported symptom severity over the course of the digital therapeutics program. In total, 89.42% of subjects experienced improvement of severity of at least one FGID. This improvement in symptom severity was not correlated with percent weight loss, gender, or age. Reduction in symptom severity was significant for all six FGIDs investigated individually and for summative symptom severity (all of them together). When we modeled the reduction in summative FGID symptom severity over the course of digital therapeutics intervention, we identified the D+G+M linear model as the best fitting, with an adjusted R^2^ of 0.442, compared with 0.124 for the D+G model and 0.318 for the D+M model. Two genomic SNPs and eight microbial taxa were the significant predictors in the best model for these participants. Thus, in our cohort, gender and baseline microbiome best classified subjects with or without FGIDs, whereas a combination of genomic SNPs and microbiome variables (but not gender) best-modeled reduction in summative FGIDs symptom severity.

We then looked at the reduction of symptom severity for three functional bowel disorders of our interest: IBS, constipation, and diarrhea. Interestingly, for IBS, the best fit model was the one containing D+G+M variables, whereas, for constipation and diarrhea, the best fit models were those only containing D+M variables. When analyzing the variables found significant to the best fit models for this cohort, we identified several genomic SNPs and microbial taxa that are shared across two or more models. SNP rs7775228 (risk allele C) was associated in the linear D+G+M model with an increase in summative symptom severity (Table 5), as well as in the linear D+G+M model with an increase in IBS symptom severity (Table 6). Interestingly, despite not being the best fit model, it also was associated in the linear D+G model with an increase in diarrhea symptom severity (Table S9) and associated in the linear D+G+M model with reduction of constipation symptom severity (Table 7). In addition to its association with gluten sensitivity [58], which is why we include this SNP as part of our care protocols, rs7775228 is involved in seasonal allergic rhinitis and as a protein biomarker for inflammation [58,59].

In terms of the microbial taxa shared across two or more models, genus *Fusicatenibacter* was associated with a greater prevalence of FGID in the D+G+M logistic model of this cohort (Table 4) and associated with an increase in IBS symptom severity in the D+G+M linear model (Table 6). Genus *Intestinimonas* was associated with a reduction in summative FGID symptom severity in the D+G+M model (Table 5) and a reduction in diarrhea symptom severity in the D+M model (Table S10). Genus *Megasphaera* was noted to be strongly associated with a reduction in summative FGID symptom severity in the D+G+M linear model of Table 5 and a reduction in IBS symptom severity in the D+G+M linear model of Table 6. Interestingly, genus *Lactobacillus* appears associated with a reduction in diarrhea symptom severity in the linear D+M model of Table S10, an effect that has been amply demonstrated in the literature [60], supporting the validity of our methods. Moreover, these bacteria, specifically, *Fusicatenibacter, Megasphaera*, and *Intestinimonas*, were either negatively associated with FGID status or were associated with a reduction in the severity of FGID symptoms are previously reported to be Short Chain Fatty Acids (SCFAs) producers [61–64]. There is ample evidence of the role of SCFAs in improving gut integrity, which plays an essential role in maintaining mucosal homeostasis [65].

Our analysis also revealed some bacterial signatures that were associated with FGID status. *Desulfovibrio* was observed to possess a significant association with an increase in FGID symptomatology (Table 5). Previous reports suggest that bacteria belonging to the genus *Desulfovibrio* generate H_2_S gas via a dissimilatory sulfate reduction pathway, leading to inflammatory gut disorders [66,67]. We also noted the association of *Akkermansia* with FGID status (Table 1). Although this bacterium is suggested to have beneficial associations with the gut health, a few studies have reported its inverse correlation with reduction of abdominal pain [68].

*Ruminococcus torques* group appears associated with FGID status in the D+G+M logistic model of Table 4 and associated with a reduction in summative FGID symptom severity in the D+G+M model of Table 5. Despite not being the best fit model, this taxon also was associated with a reduction of diarrhea symptom severity in the linear D+G+M model of Table 8. Different *Ruminococcus torques* subgroups have been associated in the literature with IBS-D, IBS-M, and Crohn’s disease subjects [57]. Additionally, genus *Terrisporobacter*, associated with inflammation and gut dysbiosis [69], was associated with FGID status in the D+G+M logistic model (Table 4). Collectively, these findings indicate a potential of gut microbial profiling not only for predicting current gastrointestinal health but also education in FGID related symptoms.

This study has some limitations that are important to note. First, the descriptive modeling exercise performed in this work is the best fit for the cohort analyzed here and is not intended to infer for the larger population. In particular, we aimed to investigate which demographic, genomic, and baseline microbiome predictors improved the fit of the models, along with the magnitude and direction of their association with FGID. Second, whereas we used all microbiome taxa present in the baseline gut microbiome samples (n=105 after the filters imposed), for genomic SNPs, we selected only markers that are used to inform diet and lifestyle interventions of subjects under the Digbi Health program, specifically those associated with intolerances and allergies (n=14). So, the fact that microbiome markers almost always outperformed genomic SNP markers may be due to the markedly different dataset sizes. Third, inclusion criteria in this study did not consider factors known to influence the microbiome composition (probiotic or antibiotic usage) or other comorbidities (musculoskeletal pain, skin conditions, hypothyroidism, diabetes, cholesterol, hypertension, and mental health) that may confound the results presented. And fourth, the survey instrument utilized was an ad-hoc questionnaire that asked participants to rate their symptom severity for different FGIDs on a scale of 1 to 5 and was not a validated clinical instrument. The survey was performed retrospectively for both time points after successfully achieving 5% or more body weight loss.

Despite the above limitations, the digital therapeutics care provided to subjects, informed by genetic and baseline gut microbiome and their interaction with participant’s lifestyle, seemed to effectively reduce symptom severity of FGIDs, including IBS, diarrhea, and constipation. Our earlier study supported the use of this care as a therapy for insulin resistance [70], empowering subjects to manage their inflammation by awareness of the impact of processed foods and foods to which they are sensitive as per their genomic SNPs and microbiome results. Dietary fiber coaching also results in increased vegetable diversity and quantity. Whereas further research is required to better understand the effect of different components of the care (e.g., fiber types) on modulating the microbial taxa and genomic SNPs identified in the models and their corresponding effect on reduction of FGIDs symptom severity, this preliminary retrospective study generates testable hypotheses for associations of a number of biomarkers in the prognosis of FGIDs symptomatology. Moreover, the methods presented add to the existing set of tools (e.g. [71]) that can be readily implemented to understand the role that genetics and gut microbiome play on disease etiology.

## Data Availability

The microbiome sequence data used in this study were submitted to NCBI SRA under accession number PRJNA760529. Additional information on subjects including demographics, weight loss, reduction in symptomatology, and genetic risk scores are provided as an electronic supplementary file.

https://www.ncbi.nlm.nih.gov/bioproject/PRJNA760529/

## Authors’ statements

### Ethics statement

E&I Review Services, an independent institutional review board, reviewed and approved IRB Study #18053 on 05/22/2018. Additionally, IRB Study #21141 was determined to be exempt from E&I Review Services on 08/06/2021. Research material derived from human participants included self-collected saliva samples using buccal swabs and fecal samples using fecal swabs. Informed consent was obtained electronically from study participants.

### Authors contribution statement

**Shreyas V Kumbhare:** Formal analysis, Methodology, Software, Visualization, Writing - original draft, Writing - review & editing; **Patricia A Francis-Lyon:** Conceptualization, Data curation, Formal analysis, Methodology, Software, Validation, Visualization, Writing - original draft, Writing - review & editing; **Dashyanng Kachru:** Data curation, Formal analysis, Methodology, Software, Visualization, Writing - original draft; **Tejaswini Uday:** Data curation, Software; **Carmel Irudayanathan:** Writing - original draft, Writing - review & editing; **Karthik M Muthukumar:** Data curation, Software; **Roshni R Ricchetti:** Writing - original draft, Writing - review & editing; **Simitha Singh-Rambiritch:** Project administration, Writing - original draft, Writing - review & editing; **Juan A Ugalde:** Software, Writing- reviewing; **Parambir S Dulai:** Writing - review & editing**; Daniel E Almonacid:** Project administration, Writing - original draft, Writing - review & editing; **Ranjan Sinha:** Conceptualization, Funding acquisition, Writing - review & editing.

## Acknowledgements

The authors would like to acknowledge Mr. Santosh Saravanan for his data mining and curation efforts.

## Supplementary Methods

### Digbi Health Program

Digbi Health is a next-generation, prescription-grade, digital therapeutics platform that analyzes genetics, gut bacteria, lifestyle, socioeconomic and behavioral risk patterns to build evidence-based individualized dietary plans, fitness and stress management programs using artificial intelligence (AI) to help reduce weight and reverse weight-related inflammatory gut, musculoskeletal, cardiovascular, and insulin-related illnesses. The digital precision care interventions are delivered via mobile app to expand the accessibility, safety, and effectiveness of health care. Digbi Health’s DNA and gut microbiome-based health program is geared primarily toward individuals who suffer from inflammatory health conditions typically associated with being overweight or obese. A large California-based health insurance payor currently covers the program for its qualifying members through its wellness platform.

Upon enrollment, Digbi Health program participants were provided with online login access to the Digbi Health app and were asked to complete a medical health questionnaire. A Bluetooth compatible digital weighing scale and saliva and stool bio sampling kits were shipped to all participating members. The App was used to track a member’s weight (via the Bluetooth scale), assess dietary intake (via uploaded photographs of food items consumed), and track wellness and lifestyle associated metrics such as sleep quality and quantity, exercise type and duration, stress and meditation, energy levels, cravings, and recommended foods consumed/avoided.

### Reports

The results presented in the genetics section of the report were determined by the number of markers and risk genotypes present in the raw genomic data, and the Digbi Health reports were loaded into the app. The Digbi Health initiative uses gut microbiome profiles (acquired via stool swab samples) to direct the course of subjects’ precision treatment, in addition to using genetic risk profiles to guide the course of therapy. Based on an analysis of these genetic and gut microbiome risk profiles, the Digbi Health Total Wellness Report was generated. The results were evaluated with the participants one-on-one by the health coach over the course of four months at pre-determined weekly and bi-weekly intervals.

### Lifestyle

The Digbi Health program is a program that uses body metrics, gut microbiome, and genetic profiles, and personalized health coaching to manage weight loss. Participants use the Digbi Health app to track ten key indicators of a healthy lifestyle and well-being (weight, sleep, hunger, stress, meditation, cravings, superfoods, morning energy, exercise and foods to avoid). They also take photos of the food they consume and are assigned a health coach who works personally with the participant through guided sessions as scheduled by the subjects to interpret the personalized wellness reports generated from participants’ app usage and a participant’s DNA and gut microbiota. The analyses also break down obesity risk depending on genetic and gut microbiota profiles of individuals. The goal of the program is for individuals to lose at least 5% of their baseline body weight by the fourth month. To achieve this goal, the program seeks to nudge participants towards making incremental lifestyle changes focused on reducing sugar consumption, timing meals to optimize insulin sensitivity, reducing systemic inflammation by identifying possibly inflammatory and anti-inflammatory nutrients via genetic testing. This is accomplished by establishing a baseline level of physical activity and maintaining it to reduce inflammation, optimizing gut health based on microbiome testing, and, most importantly, implementing these behavioral changes with the help of health coaching and the app to ensure that the changes are long-term sustainable.

## Supplementary tables

**Table S1.**
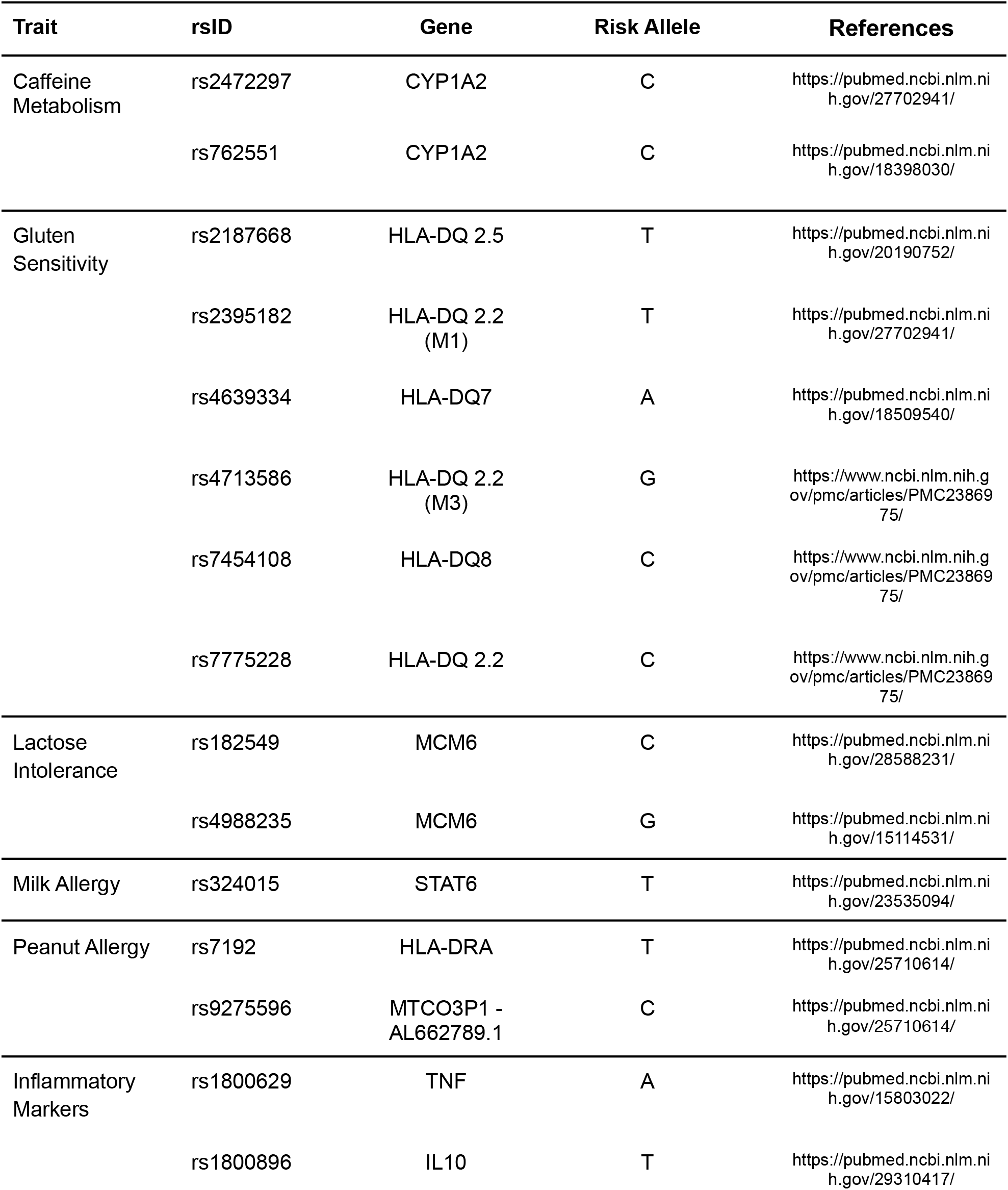
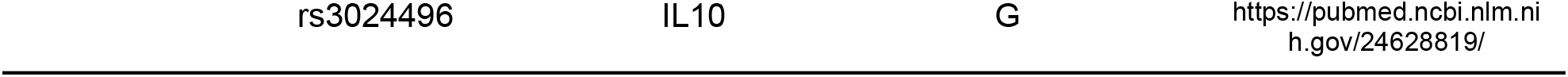
Genome SNPs utilized in this work.

**Table S2.** PERMANOVA analysis for baseline gut microbiome of subjects considering FGID status, BMI, gender, and interactions FGID status:BMI and FGID status:gender as covariates.

|  | Df | Sums Of Sqs | Mean Sqs | F. Model | R<sup>2</sup> | Pr(>F) |
| --- | --- | --- | --- | --- | --- | --- |
| FGID status | 1 | 0.00826 | 0.0082617 | 0.88650 | 0.00534 | 0.532 |
| BMI | 1 | 0.01056 | 0.0105565 | 1.13274 | 0.00682 | 0.279 |
| Gender | 1 | 0.01855 | 0.0185488 | 1.99034 | 0.01198 | 0.030* |
| FGID status:BMI | 1 | 0.00610 | 0.0061003 | 0.65458 | 0.00394 | 0.819 |
| FGID status:Gender | 1 | 0.01387 | 0.0138673 | 1.48801 | 0.00896 | 0.102 |
| Residuals | 160 | 1.49111 | 0.0093194 |  | 0.96297 |  |
| Total | 165 | 1.54844 |  |  | 1.00000 |  |

**Table S3.** Reduction in Summative FGID Symptom Severity Linear Model: Demographics + Genomics (D+G)

| Variable | Estimate | Std. Error | t value | Pr(>\|z\|) |
| --- | --- | --- | --- | --- |
| Gluten Sensitivity (rs4639334), Risk Allele A | -2.839 | 0.896 | -3.167 | 0.002* |
| Gluten Sensitivity (rs7775228), Risk Allele C | -2.763 | 0.975 | -2.835 | 0.006* |

Adjusted R<sup>2</sup>: 0.124

**Table S4.** Reduction in Summative FGID Symptom Severity Linear Model: Demographics + Microbiome (D+M)

| Variable | Estimate | Std. Error | t value | Pr(>\|z\|) |
| --- | --- | --- | --- | --- |
| *Ruminococcus torques* group | 0.727 | 0.250 | 2.915 | 0.004* |
| *Candidatus Soleaferrea* | -0.359 | 0.146 | -2.463 | 0.016* |
| *Intestinimonas* | 0.340 | 0.120 | 2.843 | 0.006* |
| Unclassified genus GCA-900066575<br>of *Lachnospiraceae* family | 0.474 | 0.143 | 3.307 | 0.001* |
| *Eubacterium hallii* group | -0.366 | 0.137 | -2.667 | 0.009* |
| *Alistipes* | -0.358 | 0.139 | -2.578 | 0.012* |
| *Megasphaera* | 0.415 | 0.132 | 3.154 | 0.002* |
| *Desulfovibrio* | -0.395 | 0.114 | -3.461 | 0.001* |

Adjusted R<sup>2</sup>: 0.318

**Table S5.** Reduction in IBS Symptom Severity Linear Model: Demographics + Genomics (D+G)

| Variable | Estimate | Std. Error | t value | Pr(>\|z\|) |
| --- | --- | --- | --- | --- |
| Gluten Sensitivity (rs7775228),<br>Risk Allele C | -0.829 | 0.276 | -3.009 | 0.004* |

Adjusted R<sup>2</sup>: 0.130

**Table S6.**
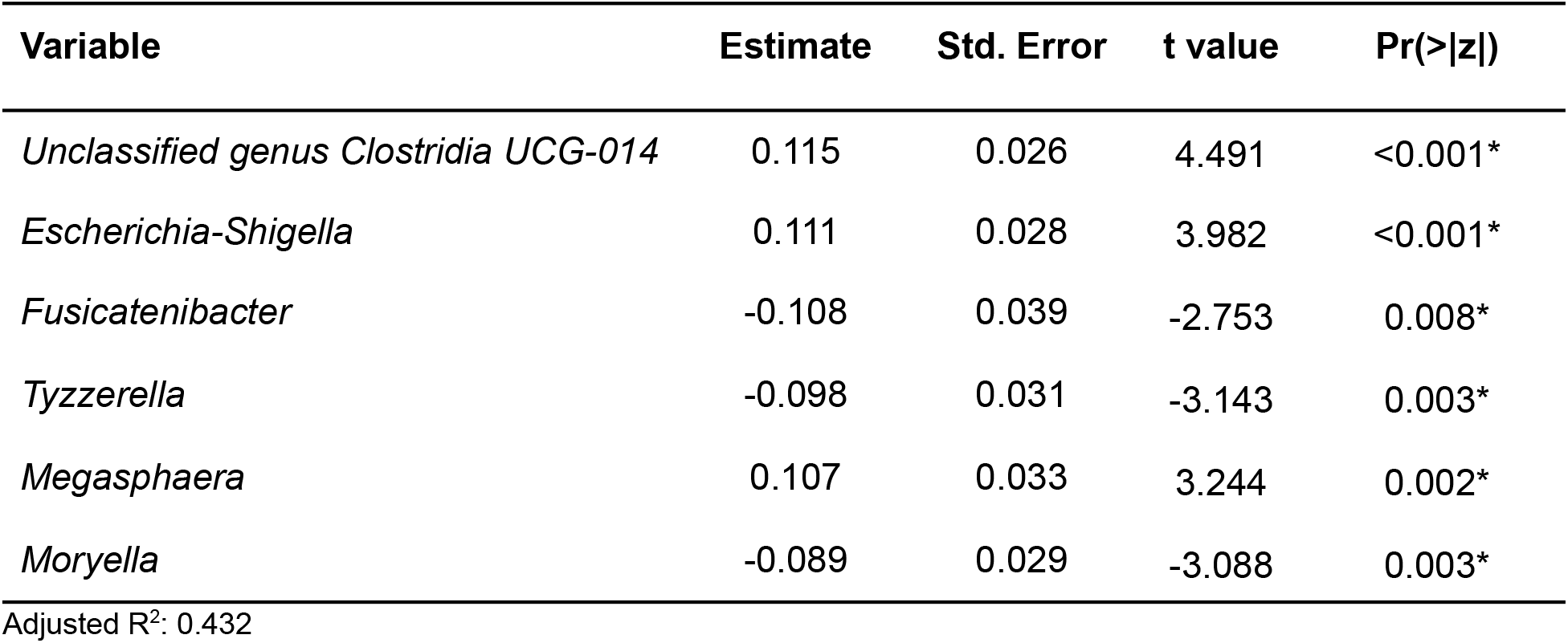
Reduction in IBS Symptom Severity Linear Model: Demographics + Microbiome (D+M)

**Table S7.** Reduction in Constipation Symptom Severity Linear Model: Demographics + Genomics (D+G)

| Variable | Estimate | Std. Error | t value | Pr(>\|z\|) |
| --- | --- | --- | --- | --- |
| Gluten Sensitivity (rs4639334), Risk Allele A | -0.558 | 0.286 | -1.948 | 0.056. |

Adjusted R<sup>2</sup>: 0.038

**Table S8.**
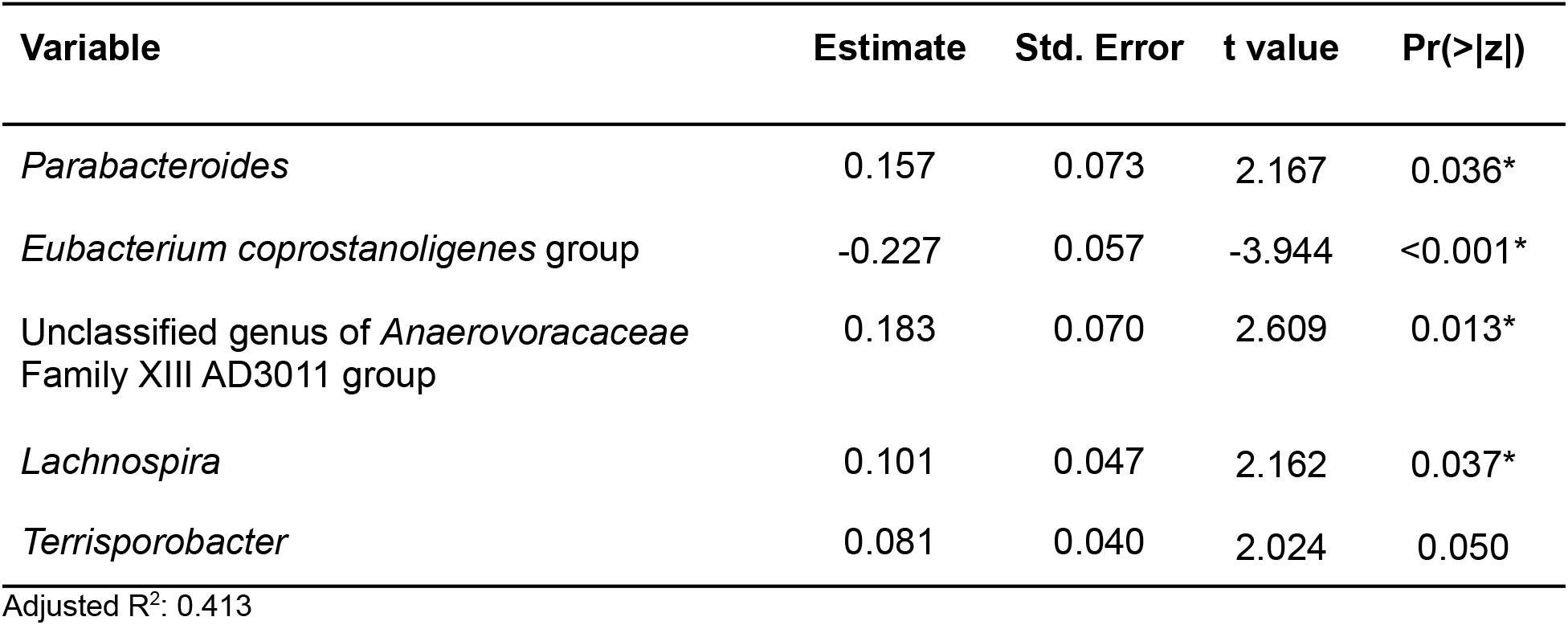
Reduction in Constipation Symptom Severity Linear Model: Demographics + Microbiome (D+M)

| Variable | Estimate | Std. Error | t value | Pr(>\|z\|) |
| --- | --- | --- | --- | --- |
| *Parabacteroides* | 0.157 | 0.073 | 2.167 | 0.036* |
| *Eubacterium coprostanoligenes* group | -0.227 | 0.057 | -3.944 | <0.001* |
| Unclassified genus of *Anaerovoracaceae*<br>Family XIII AD3011 group | 0.183 | 0.070 | 2.609 | 0.013* |
| *Lachnospira* | 0.101 | 0.047 | 2.162 | 0.037* |
| *Terrisporobacter* | 0.081 | 0.040 | 2.024 | 0.050 |

Adjusted R<sup>2</sup>: 0.413

**Table S9.** Reduction in Diarrhea Symptom Severity Linear Model: Demographics + Genomics (D+G)

| Variable | Estimate | Std. Error | t value | Pr(>\|z\|) |
| --- | --- | --- | --- | --- |
| Gluten Sensitivity (rs7775228), Risk Allele C | -0.773 | 0.330 | -2.339 | 0.024* |

Adjusted R<sup>2</sup>: 0.090

**Table S10.**
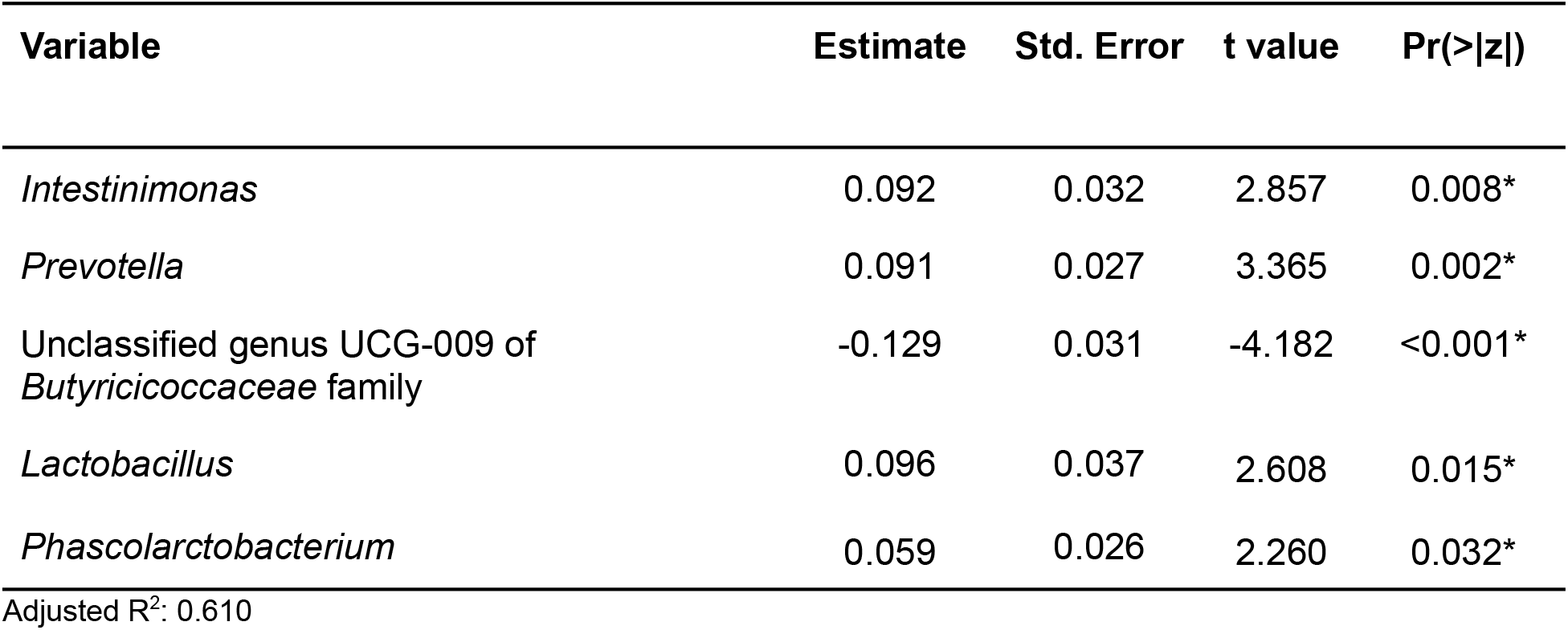
Reduction in Diarrhea Symptom Severity Linear Model: Demographics + Microbiome (D+M)

